# Comparison of bicarbonate-based versus heparin-based Impella® purge solution

**DOI:** 10.1101/2025.07.31.25332547

**Authors:** Bridget A. Betts, Miranda J. Moser, Ju Hee Kim, Danielle M. Knowles, Edmond J. Solomon, Andrew Tom, Alexander Y. Toyoda, Brian R. Schuler

## Abstract

**Background:** The Impella® is a continuous axial flow pump that utilizes a purge solution to create a positive pressure barrier and prevent pump thrombosis. Both the bicarbonate-based (BBPS) and heparin-based purge solutions (HBPS) are FDA-approved, but solution choice varies in clinical practice. This study aims to compare the efficacy and safety of the BBPS versus HBPS when BBPS is used as the standard of care.

**Methods:** This study was a retrospective analysis at two tertiary academic medical centers within the same health-system. Adult patients were included if they required an Impella® device and received either the BBPS or HBPS for at least 72 hours between July 2022 and October 2023. The major endpoint was a composite outcome of pump thrombosis, stroke, and bleeding events defined by International Society on Thrombosis and Haemostasis criteria. Pump thrombosis was defined as incidence of two or more purge pressures greater than 800 mmHg, the use of thrombolytic purge solution, or thrombus noted on imaging. Minor endpoints included intensive care unit (ICU) mortality, ICU length of stay, and vascular complications.

**Results:** A total of 178 patients were evaluated of which 66 were included. The composite outcome occurred in 23 of 38 patients (60.5%) in the BBPS group and 17 of 28 patients (60.7%) in the HBPS group (p=0.99). There was no difference in pump thrombosis, stroke or bleeding events between cohorts. Vascular events were noted in 5 (17.9%) of HBPS and 13 (34.2%) of BBPS group (p=0.14). Additional minor endpoints were not significantly different between the groups.

**Conclusions:** This study found no differences in pump thrombosis, stroke, and bleeding events in the BBPS versus HBPS group. Although these findings are supportive of utilizing BBPS as standard of care, larger, multi-center studies are needed to confirm differences in bleeding or thrombotic outcomes.

## INTRODUCTION

Cardiogenic shock is a life-threatening condition with an inpatient mortality rate up to 51%.^1^ Patients in cardiogenic shock often require inotropic support and, in refractory cases, temporary mechanical circulatory support (MCS) devices such as the Impella® to maintain adequate cardiac output. Although these devices may be lifesaving when medical therapy fails, they introduce potential complications such as bleeding, hemolysis, device thrombosis, vascular and valvular injury, and arrhythmias.^2^ To lower the risk of device thrombosis and prolong motor life, the Impella® uses a countercurrent flow purge solution that creates a pressure barrier that prevents blood from entering the pump motor.^3^

ABIOMED® (Danvers, MA) recommends a heparin-based (25 or 50 U/mL) purge solution (HBPS), along with systemic intravenous (IV) heparin for anticoagulation to reduce the risk of thrombotic complications.^2^ The HBPS introduces an additional layer of complexity in managing systemic anticoagulation due to multiple sources of heparin leading to variable amounts of heparin being infused each hour^4^.ABIOMED® recommends the use of sodium bicarbonate-based (25 or 50 mEq/L) purge solutions (BBPS) as an alternative to heparin-based purge solutions for patients intolerant to heparin or in whom heparin is contraindicated. Sodium bicarbonate protects against device thrombosis by increasing protein stability and reducing protein denaturation without interfering with the coagulation cascade.^5^

The data supporting the safety and efficacy of BBPS is limited. Some in vitro data and small retrospective studies are available, but data evaluating patient-based bleeding and thrombosis outcomes is scarse.^6^ In 2022, Massachusetts General Brigham enterprise (MGB) changed their clinical practice to utilize a BBPS as the standard purge solution in all patients who have an Impella® device placed instead of a HBPS. With the limited data that exists for the use of BBPS, our study aims to compare the thrombosis and bleeding rates in patients receiving Impella® support with a HBPS compared to a BBPS.

## MATERIALS AND METHODS

### Design

This analysis was a retrospective, multicenter cohort study including patients at Massachusetts General Hospital (MGH) or Brigham and Women’s Hospital (BWH) who were supported with an Impella® device pre (January 2022 – October 2022) and post (January 2023 – October 2023) utilization of BBPS as the standard of care. Patients in the “pre” group received a HBPS (25 or 50 units/mL) and patients in the “post” group received BBPS (25 or 50 mEq/L). Standardization of data collection database input entries and definitions of all collection points was pre-determined before collection was started. The protocol of this retrospective study was approved by the MGB Institutional Review Board Protocol #2023P002761.

Patient lists were generated through the electronic health record of all patients at BWH or MGH who received HBPS or BBPS orders. Adult patients who were admitted to either institution with an Impella® device placed for greater than 72-hours and received systemic anticoagulation titrated to a goal partial thromboplastin time (PTT) were included in this analysis. Patients were excluded if they received a bivalirudin purge solution, systemic heparin titrated to anti-Xa levels, had an elevated baseline partial thromboplastin time (PTT) prior to anticoagulation (PTT > 37.5 seconds), unknown baseline PTT, or received a fixed rate, non-titratable heparin infusion.

### Baseline Characteristics

Data collected included demographics, admission diagnosis, hospital length of stay, intensive care unit (ICU) length of stay, and body mass index (BMI). The following data points were collected to assess bleeding or thrombosis risk and clinical status at time of Impella® placement: previous cardiac surgery and/or intervention, past medical history, medications that are related to increased bleeding or thrombotic risk (e.g, anticoagulants, antiplatelets, and oral contraceptives), cardiac arrest on admission, extracorporeal membrane oxygenation (ECMO), and Sequential Organ Failure Assessment (SOFA) score at baseline. Impella® placement time was obtained through procedural documentation.

Baseline laboratory values were defined as the value closest to Impella® placement (within 24-hours) with preference for pre-placement, but if lab values were not available pre-placement, then a value obtained within two hours following placement was recorded as a baseline value.

### Outcomes

Our major outcome was the composite of pump thrombosis, stroke, and bleeding. Pump thrombosis was defined as the incidence of two or more purge pressures greater than 800 mmHg, use of a thrombolytic purge solution, or thrombus noted on report or imaging (e.g., echocardiogram) thought to be related to the device. The purge pressure threshold was selected based on previous literature associating a purge pressure > 800 mmHg with Impella® pump thrombosis.^2^ The major, clinically relevant non-major, and minor bleeding was defined by the International Society on Thrombosis and Haemostasis (ISTH) criteria.^8^ Minor outcomes included ICU length of stay, ICU mortality, time to first therapeutic PTT, percentage of PTTs within the therapeutic range during the first 72-hours, incidence of supratherapeutic PTTs during impella insertion, vascular complications as defined by Valve Academic Research Consortium-2 criteria, number of blood products transfused, and incidence of heparin-induced thrombocytopenia.^9^ The percentage of time in therapeutic range was calculated based on values collected in concordance with the goals at that time. Variations of those goals based on patient factors were also collected.

### Statistical Analysis

Descriptive data was used to describe the populations. Categorical data was assessed using Chi-squared tests. Nonparametric continuous variables were assessed using Mann-Whitney U test and Fisher’s exact test as appropriate.

## RESULTS

### Patient Population

A total of 178 patients who received an Impella® device were evaluated for inclusion between the two academic medical centers of which 66 patients were included (44 patients from MGH and 22 patients from BWH). The most common reason for exclusion was Impella® support for < 72 hours (Figure 1). Of the 66 patients included, 38 received a BBPS and 28 patients received a HBPS. The median age was 62.5 and 59.5 years old in the HBPS and BBPS groups, respectively, and the primary indication for Impella® placement was cardiogenic shock. The most common device inserted was the Impella® 5.5. The median duration of Impella® placement was 7.5 days in the HBPS group and 8.3 days in the BBPS group. Additional relevant baseline characteristics and pertinent comorbidities can be found in Tables 1-3.

**Table 1.** Baseline Characteristics.

|  | Heparin-based purge solution group (n=28) | Bicarbonate-based purge solution group (n= 38) |
| --- | --- | --- |
| Age, years <sup>a</sup> | 62.5 (53.8 – 68.3) | 59.5 (47.3 – 68.8) |
| Sex <sup>b</sup> |  |  |
| Male | 23 (82.1) | 26 (68.4) |
| Race <sup>b</sup> |  |  |
| White | 23 (82.1) | 28 (73.7) |
| Black | 3 (10.7) | 6 (15.8) |
| Unknown | 2 (7.2) | 3 (7.9) |
| Asian | 0 | 1 (2.6) |
| Body Mass Index , kg/m<sup>2</sup> <sup>a</sup> | 26.4 (23.1 – 28.8) | 26.1 (23.5 – 29.3) |
| Relevant Comorbidities <sup>a</sup> |  |  |
| CHF | 17 (60.7) | 22 (57.9) |
| CAD | 17 (60.7) | 18 (47.4) |
| Hypertension | 16 (57.1) | 23 (60.5) |
| Diabetes | 12 (42.9) | 14 (36.8) |
| CKD | 9 (32.1) | 12 (31.6) |
| Current or former smoker | 8 (28.6) | 4 (10.5) |
| Atrial fibrillation | 5 (17.9) | 13 (34.2) |
| History of thrombosis (stroke, PE, DVT) | 4 (14.3) | 6 (15.8) |
| PAD/PVD | 3 (10.7) | 1 (2.6) |
| History of hypercoagulability | 1 (3.6) | 3 (7.9) |
| History of mitral stenosis | 1 (3.6) | 1 (2.6) |
| History of cirrhosis | 0 | 1 (2.6) |
| History of HIT | 0 | 2 (5.2) |
| Malignancy | 0 | 3 (7.9) |
| <sup>a</sup>Data presented as median (IQR); <sup>b</sup> Data expressed as n (%)<br>CAD: coronary artery disease; CHF: congestive heart failure; CKD: chronic kidney disease; HIT: heparin-induced thrombocytopenia; PE: pulmonary embolism; DVT: deep vein thrombosis; PAD: peripheral artery disease; PVD: peripheral vascular disease |  |  |

**Table 2.** Baseline Characteristics related to Impella® support.

|  | Heparin-based purge solution group (n=28) | Bicarbonate-based purge solution group (n=38) |
| --- | --- | --- |
| Device type <sup>a</sup> |  |  |
| Impella® 5.5 | 17 (60.7) | 27 (71) |
| Impella® CP | 11 (39.3) | 9 (23.7) |
| Indication for device<sup>a</sup> |  |  |
| Cardiogenic shock | 24 (85.7) | 33 (86.8) |
| Procedure | 4 (14.3) | 5 (13.2) |
| Heparin given during placement, units<sup>b</sup> | 11000 (6250 – 18625) | 10000 (7375 – 14875) |
| Duration of Impella® placement, days<sup>b</sup> | 7.5 (5.2 – 12.2) | 8.3 (6.2 – 14.3) |
| Cardiac arrest on admission<sup>a</sup> | 2 (7.1) | 5 (13.2) |
| Extracorporeal Membrane Oxygenation<sup>a</sup> | 9 (32.1) | 10 (26.3) |
| Relevant baseline medications<sup>a</sup> |  |  |
| **Anticoagulants** |  |  |
| Anti-Xa inhibitor | 1 (3.6) | 1 (2.6) |
| Bivalirudin | 1 (3.6) | 4 (10.5) |
| **Antiplatelets** |  |  |
| Aspirin | 3 (10.7) | 1 (2.6) |
| Cangrelor | 3 (10.7) | 2 (5.3) |
| Oral P2Y12 inhibitor | 1 (3.6) | 1 (2.6) |
| <sup>a</sup> Data expressed as n (%); <sup>b</sup> Data presented as median (IQR) |  |  |

**Table 3.** Additional baseline characteristics.

|  | Heparin-based purge solution group (n=28) | Bicarbonate-based purge solution group (n=38) | p-value |
| --- | --- | --- | --- |
| Sequential Organ Failure Assessment (SOFA) score | 6.5 (4.8 – 10) | 7 (5 – 8.75) | 0.93 |
| Lactate, (mmol/L)<sup>b</sup> | 2.4 (1.4 – 5) | 2 (1.3 – 3.8) | 0.36 |
| Hemoglobin, (g/dL) | 10.2 (8.8 – 13) | 10.2 (8.5 – 12.6) | 0.47 |
| <sup>a</sup> Data presented as median (IQR) <sup>b</sup> n=27 |  |  |  |

**Figure 1.**
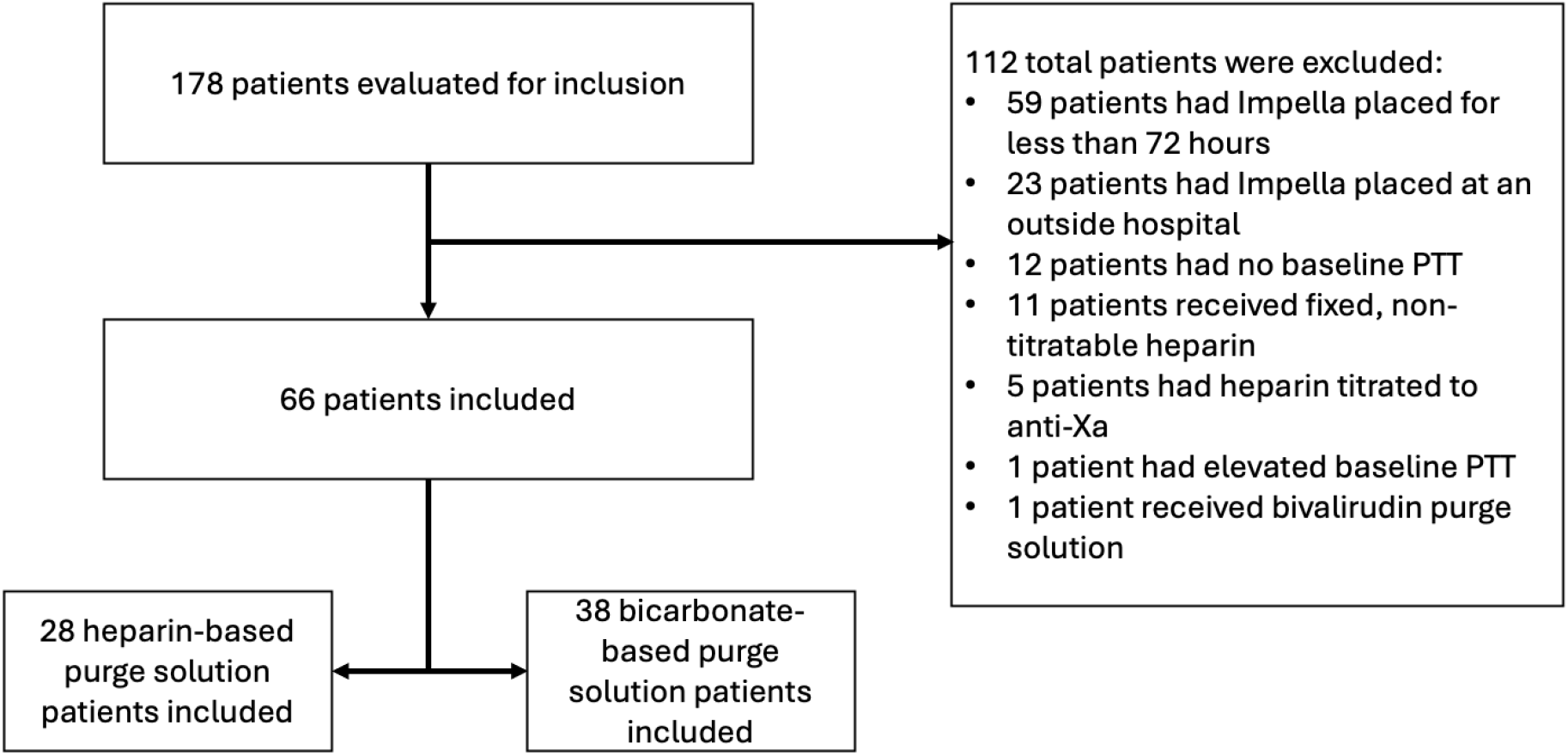
Screening of patient population.

### Major Outcome

Our major composite outcome of pump thrombosis, stroke, or bleeding occurred in 23 of 38 patients (60.5%) in the BBPS group and 17 of 28 patients (60.7%) in the HBPS group (p=0.99) (Table 4). The incidence of pump thrombotic events was 23.7% vs.17.9% (p=0.57), the incidence of stroke was 13.2% vs. 7.1% (p=0.69) and the rate of bleeding events was 25% vs. 35.7% in (p=0.29) the BBPS vs. HBPS group, respectively (Table 4).

**Table 4.**
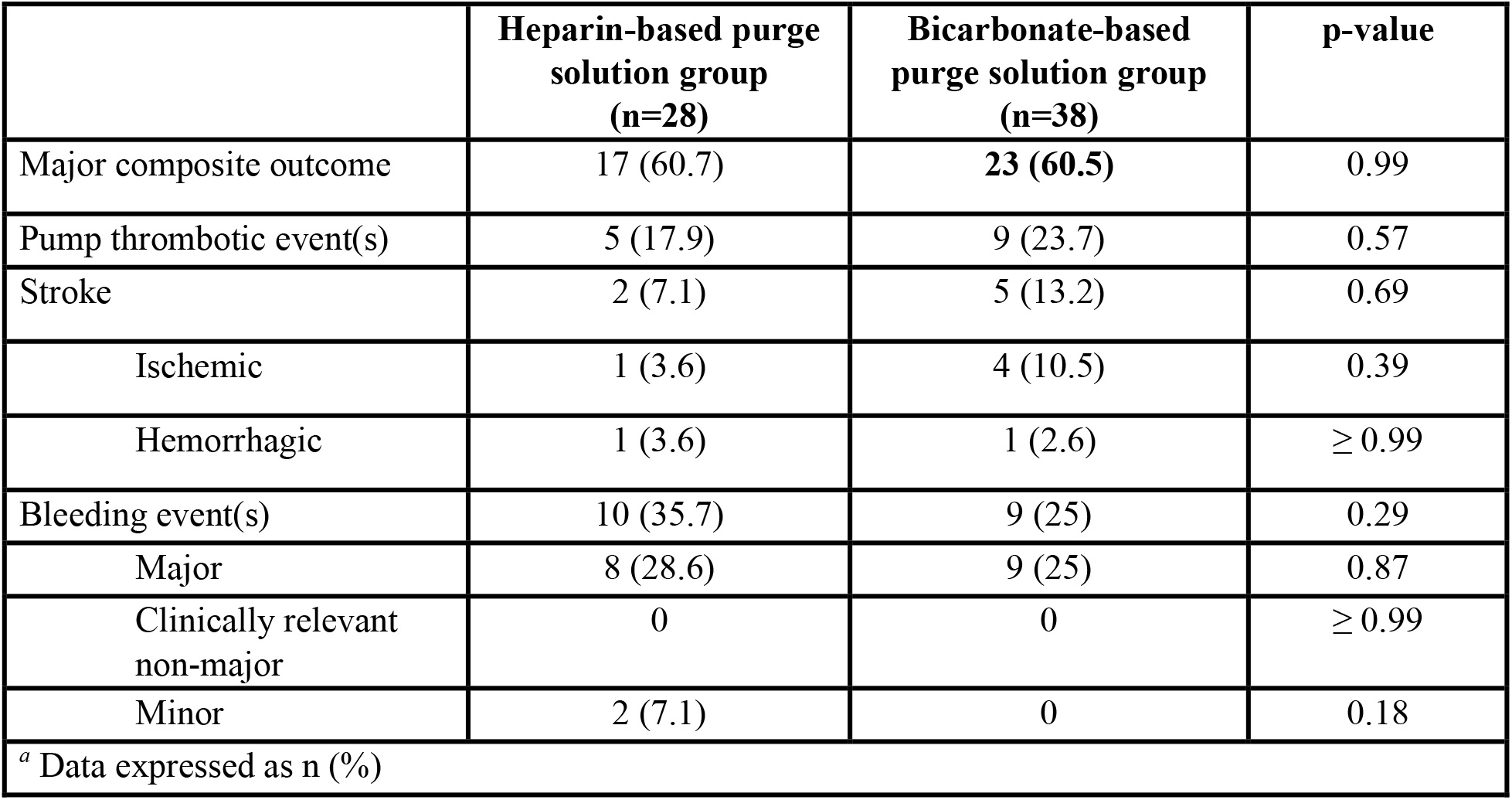
Major Composite Outcome.

|  | Heparin-based purge solution group (n=28) | Bicarbonate-based purge solution group (n=38) | p-value |
| --- | --- | --- | --- |
| Major composite outcome | 17 (60.7) | **23 (60.5)** | 0.99 |
| Pump thrombotic event(s) | 5 (17.9) | 9 (23.7) | 0.57 |
| Stroke | 2 (7.1) | 5 (13.2) | 0.69 |
| Ischemic | 1 (3.6) | 4 (10.5) | 0.39 |
| Hemorrhagic | 1 (3.6) | 1 (2.6) | ≥ 0.99 |
| Bleeding event(s) | 10 (35.7) | 9 (25) | 0.29 |
| Major | 8 (28.6) | 9 (25) | 0.87 |
| Clinically relevant non-major | 0 | 0 | ≥ 0.99 |
| Minor | 2 (7.1) | 0 | 0.18 |
| <sup>a</sup> Data expressed as n (%) |  |  |  |

### Minor Outcomes

Results for the minor outcomes can be found in Table 5. The rate of vascular events was 34.2% in the BBPS group and 17.9% in the HBPS group (p=0.14). The percentage of PTTs within the therapeutic range during the first 72-hours of Impella® placement were similar (41% vs 41%); however, there was a nonsignificant trend toward a longer duration of time to achieve a first therapeutic PTT in the HBPS group (24.6 vs 27.7 hours).

**Table 5.** Minor Outcomes.

|  | Heparin-based purge solution group (n=28) | Bicarbonate-based purge solution group (n=38) | p-value |
| --- | --- | --- | --- |
| Vascular event(s) <sup>a</sup> | 5 (17.9) | 13 (34.2) | 0.14 |
| Major | 1 (3.6) | 4 (10.5) | 0.29 |
| Minor | 4 (14.3) | 9 (25) | 0.34 |
| Total length of stay, days <sup>b</sup> | 26.5 (15.8 – 43.8) | 32.5 (21.5 – 47.5) | 0.24 |
| Length of ICU admission, days <sup>b</sup> | 16.5 (10.5 – 35) | 20 (13.3 – 32) | 0.33 |
| ICU mortality <sup>a</sup> | 10 (35.7) | 11 (28.9) | 0.56 |
| PTTs within therapeutic range during the first 72-hours of placement, % <sup>b</sup> | 41 (23 – 53) | 41 (30 – 50) | ≥0.99 |
| Time to first therapeutic PTT, hours <sup>b</sup> | 27.7 (13.1 – 46.3) | 24.6 (13.4 – 31.6)<br>*n=37* | 0.21 |
| Incidence of supratherapeutic PTTs <sup>a</sup> | 8 (28.6) | 12 (31.6) | 0.79 |
| Total blood products transfused during the first 24-hours post-Impella® placement, units |  |  |  |
| pRBC | 38 | 33 | -- |
| FFP | 11 | 16 | -- |
| Platelets | 9 | 5 | -- |
| Cryo | 1 | 1 | -- |
| Total blood products transfused after 24-hours up to Impella® removal, units |  |  |  |
| pRBC | 117 | 101 | -- |
| FFP | 19 | 14 | -- |
| Platelets | 44 | 13 | -- |
| Cryo | 5 | 4 | -- |
| Number of patients that required transfusions <sup>a</sup> |  |  |  |
| pRBC | 18 (64.3) | 22 (57.9) | -- |
| FFP | 5 (17.9) | 9 (23.7) | -- |
| Platelets | 7 (25) | 9 (23.7) | -- |
| Cryo | 1 (3.6) | 2 (5.3) | -- |
| Average blood products transfused during the first 24-hours post-Impella® placement per patient, units |  |  |  |
| pRBC | 2.1 | 1.5 | -- |
| FFP | 2.2 | 1.8 | -- |
| Platelets | 1.3 | 0.5 | -- |
| Cryo | 1 | 0.5 | -- |
| Average blood products transfused after 24-hours up to Impella® removal per patient, units |  |  |  |
| pRBC | 6.5 | 4.6 | -- |
| FFP | 3.8 | 1.6 | -- |
| Platelets | 6.3 | 1.4 | -- |
| Cryo | 5 | 2 | -- |
| <sup>a</sup> Data expressed as n (%); <sup>b</sup> Data presented as median (IQR)<br>Cryo: Cryoprecipitated antihemophilic factor; FFP: Fresh frozen plasma; pRBC: Packed red blood cells |  |  |  |

## DISCUSSION

To date, BBPS has not been compared to HBPS in a randomized controlled trial. Despite this, our institutions have used an approach that is different than others. The rationale was that a BBPS would avoid multiple sources of heparin which could potentially help standardize anticoagulation practice and minimize bleeding. However, BBPS would also come with the potential risk of increased thrombotic complications given decreased local anticoagulant effect at the outflow site. We observed no significant difference in thrombotic or bleeding outcomes between patients who received a BBPS or a HBPS in this retrospective cohort study. Our study adds to the limited body of evidence that BBPS may offer a safe and effective alternative to HBPS for Impella® device success. Additionally, a trend towards more vascular events, primarily driven by the presence of hematoma, was seen in the BBPS group, although not statistically different.

The most common reason for patient exclusion was Impella® placement for less than 72-hours. We excluded these patients to limit events that happened early on, since the placement technique or issues related to the pump itself would likely drive our events instead of the choice of purge solution during the first 72 hours. Of patients that met inclusion criteria, the baseline characteristics were similar between the two groups. There was a numerically higher percentage of patients who had a cardiac arrest on admission in the BBPS group compared to the HBPS group (13.2% vs 7.1%). Additionally, more patients in the BBPS group had a history of atrial fibrillation (34.2% vs 17.9%) which may further explain why a numerically higher rate of pump thrombosis (9/38 vs 5/28 patients) and stroke (5/38 vs 2/28 patients) was seen in this group compared to the HBPS group. To classify the level of organ dysfunction and mortality risk in ICU patients, values incorporated into SOFA scoring were collected. Overall, the median SOFA score was 7 and 6.5 for the BBPS group and HBPS groups, respectively, suggesting a similar acuity in both groups.

In comparison to other literature, we utilized a composite outcome of the events. The utility of composite rather than previous studies looking solely on bleeding or thrombosis serves to fill a large gap in previous literature. Our study’s thorough definitions of outcomes, including validated ISTH and VARC-2 criteria encompassed a more involved clinical representation of outcomes rather than surrogate laboratory markers such a fall in hemoglobin. Both thrombotic and bleeding events were similar between the groups; however, the difference was not statistically significant. Although not statistically significant, a trend toward more bleeding events (both major and minor) occurred in the HBPS group. Our findings are comparable to another retrospective study that investigated the two purge solutions. A similar approach was taken to define pump related thrombosis with multiple purge pressures greater than 800 mmHg, thrombolytics in purge solution, and Impella® replacement.^2^ ICU length of stay and duration of Impella® placement was notably longer in the bicarbonate group, highlighting that patients in this group could be patients awaiting transplant. Strokes were numerically higher in the BBPS group and larger studies would be needed to determine if the incidence is truly different. This is an outcome of interest because of the presumed decrease in local anticoagulant effect with heparin removal from the purge solution. Our findings suggest, selection of purge solution should be patient-specific based on knowledge of risk factors.

Regarding secondary findings, a trend for more vascular events in the BBPS group was present (34.2% vs 17.9%). A frequent example of major vascular events that occurred in our patient population being access site or access-related vascular injury including hematoma. In a recent study looking at Impella® usage for infarct-related cardiogenic shock, Møller et al. found a high incidence of vascular related adverse effects (10% versus 3%) in patients that received an Impella®.^10^ This potentially prompts further study into purge solution’s role in minimizing vascular events.

The time to first therapeutic PTTs in the BBPS group was 24.6 hours versus 27.7 hours in the HBPS group. This may be due to institutional practice of empirically adjusting initial systemic heparin rates to account for the additional heparin provided by the heparin-based purge solution in the HBPS group. Each institution’s respective PTT goal was collected in coordination with the patient PTT. This data was collected for up to 72-hours to gather information that occurred in that time frame.

Limitations to our study are certainly present. First, it was a retrospective study and reliant on proper documentation in provider and nursing notes. Given the retrospective nature of the study, unclear documentation of timing events during operating room or cardiac catheterization lab procedures presented challenges. Additionally, the study period included a relatively small sample size; therefore, we could not perform a power or sample size calculation. Retrospective review of PTT goals and available PTTs documented also presented challenges in the data collection process. Strengths of our study include the multi-site design providing more generalizability across multiple practices. BBPS was utilized as standard of care at both institutions, minimizing risk of selection bias. Study definitions for vascular and bleeding events were well defined and stratified based on severity with clear, validated definitions.

## CONCLUSION

This study hypothesis generates the utility and safety of bicarbonate-based purge solutions as an alternative to the previous standard of care, heparin-based purge solutions. Bicarbonate based purge solution is currently only FDA approved for patients who are unable to tolerate heparin or have a contraindication to heparin, although this novel study highlights potential shift in practice given two institution’s experience of safe and efficacious use of bicarbonate as the standard of care. Additionally, larger, multi-center studies are needed to confirm if differences in bleeding or thrombotic outcomes exist.

## Data Availability

Data may be made available to reviewers as requested.

## Notes

No financial disclosures or conflicts of interest

### Competing Interest Statement

The authors have declared no competing interest.

### Funding Statement

No funding was received.

### Author Declarations

The protocol of this retrospective study was approved by the Mass General Brigham Institutional Review Board Protocol #2023P002761.

